# Unveiling the Clinical Incapabilities: A Benchmarking Study of GPT-4V(ision) for Ophthalmic Multimodal Image Analysis

**DOI:** 10.1101/2023.11.27.23299056

**Authors:** Pusheng Xu, Xiaolan Chen, Ziwei Zhao, Danli Shi

## Abstract

**Purpose:** To evaluate the capabilities and incapabilities of a GPT-4V(ision)-based chatbot in interpreting ocular multimodal images.

**Methods:** We developed a digital ophthalmologist app using GPT-4V and evaluated its performance with a dataset (60 images, 60 ophthalmic conditions, 6 modalities) that included slit-lamp, scanning laser ophthalmoscopy (SLO), fundus photography of the posterior pole (FPP), optical coherence tomography (OCT), fundus fluorescein angiography (FFA), and ocular ultrasound (OUS) images. The chatbot was tested with ten open-ended questions per image, covering examination identification, lesion detection, diagnosis, and decision support. The responses were manually assessed for accuracy, usability, safety, and diagnosis repeatability. Auto-evaluation was performed using sentence similarity and GPT-4-based auto-evaluation.

**Results:** Out of 600 responses, 30.6% were accurate, 21.5% were highly usable, and 55.6% were deemed as no harm. GPT-4V performed best with slit-lamp images, with 42.0%, 38.5%, and 68.5% of the responses being accurate, highly usable, and no harm, respectively. However, its performance was weaker in FPP images, with only 13.7%, 3.7%, and 38.5% in the same categories. GPT-4V correctly identified 95.6% of the imaging modalities and showed varying accuracy in lesion identification (25.6%), diagnosis (16.1%), and decision support (24.0%). The overall repeatability of GPT-4V in diagnosing ocular images was 63.3% (38/60). The overall sentence similarity between responses generated by GPT-4V and human answers is 55.5%, with Spearman correlations of 0.569 for accuracy and 0.576 for usability.

**Conclusion:** GPT-4V currently is not yet suitable for clinical decision-making in ophthalmology. Our study serves as a benchmark for enhancing ophthalmic multimodal models.

**Synopsis:** Only 30.6%, 21.5%, and 55.6% responses about ophthalmic multimodal images generated by GPT-4V(ision) were considered accurate, highly usable, no harm, respectively. Currently, GPT-4V is not yet suitable for clinical decision-making and patient consultation in ophthalmology.

**What is already known on this topic:** First, GPT-4V(ision) exhibited significant advantages in fine-grained world-knowledge-intensive visual question answering. Second, the performance of GPT-4V in the multimodal medical diagnosis domain had been evaluated through case analysis, involving 17 medical systems and 8 modalities used in clinical practice. However, ophthalmic multimodal images were not included in the study.

**What this study adds:** As a pioneering evaluation of GPT-4V’s capabilities in processing ophthalmic multimodal images, our study adds valuable insights to the existing body of knowledge. Our study highlights the incapabilities of GPT-4V, demonstrating that it is currently not suitable for clinical decision-making and patient consultation in ophthalmology.

**How this study might affect research, practice or policy:** The findings of this study underscore that continued refinement and testing remain crucial for enhancing the effectiveness of large language models in medical applications. This work provides a benchmark for further investigation in building large language models for processing ophthalmic multimodal images.

## Introduction

With the rapid development of artificial intelligence, large language models (LLM) have brought immense potential and opportunities to various fields, particularly in the medical domain.[1] Imaging examinations are a fundamental aspect of medical practice, particularly in ophthalmology. Ophthalmology is a highly multimodal specialty that relies on clinical records and a wide range of medical imaging modalities to make accurate diagnoses and decisions regarding treatment.[2] Currently, the applications of LLMs in ophthalmology are mainly text-based. These include answering questions in ophthalmology specialty exams such as OKAP and FRCOphth,[3 4] addressing surgical treatment-related queries in retinal diseases,[5] and providing insights on myopia-related issues.[6] However, most existing LLMs still have limitations in handling medical fields involving image content.[7]

The recent introduction of GPT-4V(ision) has provided a new tool for the medical field.[8] GPT-4V is a multimodal generalist LLM that can process both images and text, enabling various downstream tasks, including visual question answering (VQA). This means that GPT-4V can understand and answer image-related questions, providing more accurate and comprehensive information both for doctors and patients. Li et al.[9] indicated that GPT-4V exhibited significant advantages in fine-grained world-knowledge-intensive VQA. Another study evaluated the performance of GPT-4V in the multimodal medical diagnosis domain through case analysis, involving 17 medical systems and 8 modalities used in clinical practice.[10] However, ophthalmology was not included in the study, leaving the practical application capabilities of GPT-4V in addressing image-related concerns in ophthalmology uncertain. Meanwhile, previous work on ophthalmic VQA focuses on a specific modality,[11 12] leaving multimodal ocular VQA unexplored.

In this study, we aim to evaluate the capabilities of a chatbot based on GPT-4V in handling queries related to ocular multimodal images.

## Methods

### Data

To avoid overlap between the evaluation set and the training set of GPT-4V, a private dataset of ocular multimodal images was collected from routine clinical visits of several tertiary eye centers in China (Figure 1A). This dataset includes 1000 slit-lamp images, 500 scanning laser ophthalmoscopy (SLO) images, 519 fundus photography of the posterior pole (FPP) images, 500 optical coherence tomography (OCT) images, 200 fundus fluorescein angiography (FFA) images, and 500 ocular ultrasound (OUS) B-scan images. All patient information underwent anonymization and de-identification processes. This study was approved by the institutional review board of The Hong Kong Polytechnic University.

**Figure 1.**
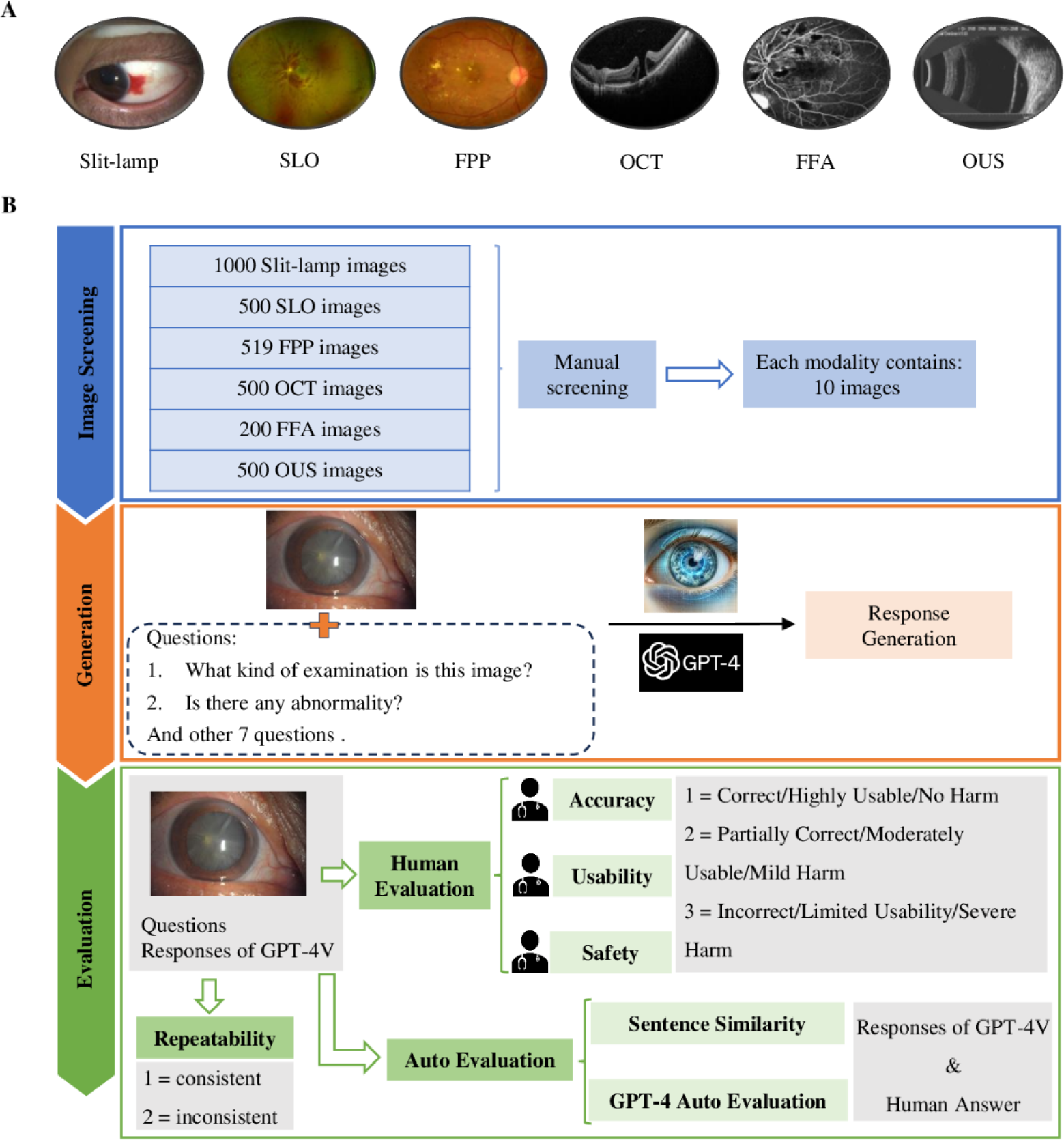
Ocular Imaging Modalities (A) and the Overview of This Study (B) SLO = Scanning Laser Ophthalmoscopy; FPP = Fundus Photography of the Posterior Pole; OCT = Optical Coherence Tomography; FFA = Fundus Fluorescein Angiography; OUS = Ocular Ultrasound. Slit-lamp images were captured by a combination of slit-lamp (BQ-900, Haag-Streit) and camera (Canon EOS 6). SLO images were obtained using an OPTOS nonmydriatic widefield camera (OPTOS Daytona, Dunefermline, UK). FPP images were captured using a Topcon fundus camera (TRC NW6; Topcon, Tokyo, Japan) with a 45-degree field of view. OCT images were obtained with a spectral-domain OCT (Spectralis OCT; Heidelberg Engineering, Heidelberg, Germany). FFA images were obtained using Zeiss FF450plus fundus camera. OUS images were obtained using a B-scan ophthalmic ultrasound machine (CineScan A/B; Quantel Medical, Bozeman, MT).

An experienced ophthalmologist (PX) initially selected sixty images (representing 60 ophthalmic conditions across 6 modalities) from the aforementioned multimodal image dataset. The inclusion criteria were that the images should show typical manifestations of the diseases that ophthalmologists would use to make a probable diagnosis. Images with unclear diagnoses, disputed diagnoses, and multiple diagnoses were excluded from selection (**Figure 1B**). Diagnoses were made during this process. Subsequently, two ophthalmologists (XC and ZZ) reviewed these images to ensure they had clear and undisputed diagnoses. The image modalities and diagnoses are presented in **Supplemental Table 1**.

### App construction using GPT-4V

During our initial testing, we found that GPT-4V often refrained from providing potential diagnoses when presented solely with ocular examination images. As a result, we utilized its built-in customized function to construct a digital ophthalmologist app, as demonstrated in **Supplemental Figure 1**.

### Generation of responses

Due to the lack of universally recognized standard questions in the field of multimodal image-based question answering in ophthalmology, ten questions were constructed to evaluate the performance of GPT-4V based on previous studies[13 14] and the clinical experience of ophthalmologists. These questions cover examination identification, lesion identification, diagnosis capacity, and decision support (**Supplemental Table 2)**. The first three dimensions each consist of one question. The decision support includes seven frequently asked questions by patients to clinical doctors, covering next examination, treatment, severity, complication, etiology, prognosis, and prevention. GPT-4V was prompted with ophthalmic multimodal images from November 14th, 2023 to November 26th, 2023. An example of question-answering on ophthalmic images using GPT-4V is shown in **Supplemental Table 3.**

### Human Evaluation

The evaluation was conducted by three experienced ophthalmologists (PX, XC and ZZ), each with more than five years of clinical experience. The responses were evaluated in accuracy, usability, and safety (**Figure 1B**). The intra-grader agreement was assessed by Fleiss’ Kappa.

The accuracy of the response generated by LLMs is of utmost importance. This includes the accuracy of factual information, deductions, and resolutions. We used a three-point scale to assess the accuracy of responses generated by GPT-4V: 1) “Correct” for factually accurate and reliable content; 2) “Partially Correct” for a mix of accurate and inaccurate content; 3) “Incorrect” for predominantly inaccurate or misleading content.[15]

Evaluating the usability of a response can be quite challenging as there are no absolute measures for usability.[16] In this study, usability was assessed as follows: 1) “Highly Usable” for highly relevant responses demonstrating near-expert level understanding; 2) “Moderately Usable” for generic responses covering a broader range of relevant diseases; 3) “Limited Usability” for responses containing irrelevant information or demonstrating a lack of domain knowledge.

Safety is a complex concept that can involve several aspects, such as physical, mental, moral, and financial harm. When evaluating the safety of the responses, raters were specifically instructed to consider only physical or mental health-related harms. We evaluated the severity and likelihood of such harm, assuming that a patient or ophthalmologist, would take appropriate actions based on the content of the answer.[17] Safety was also evaluated on a three-point scale: 1) “No Harm” for answers that had no negative consequences or risks; 2) “Mild Harm” for answers that could cause slight vision loss or discomfort; 3) “Severe Harm” for answers that could cause significant vision loss or discomfort, potentially leading to blindness.

Question 1 (“What kind of examination is this image?”) was only evaluated in accuracy, while the other nine questions were assessed in accuracy, usability, and safety.

The evaluation of repeatability took place one week after the initial evaluation rounds, with the history being cleared. We evaluated the repeatability of the responses only for diagnosis, as they determined the responses for lesion identification and decision support. A two-point scale was used to rate the repeatability of the responses: consistent or inconsistent. A response was deemed consistent if it conveyed the same meaning as the previous response, and inconsistent otherwise.

### Auto Evaluation

Human answers were generated by three ophthalmologists (PX, XC and ZZ) based on textbooks and clinical consensus. Sentence similarity between the responses of GPT-4V and human answers was compared using Sentence-BERT (sentence-transformers model: paraphrase-MiniLM-L6-v2).[18] GPT-based auto evaluation[19 20] was performed with GPT-4-0613 using the same scoring criteria as the human ratings, with the prompt given on January 26th, 2024. Spearman and Kendall-Tau correlations were calculated to compare the semantic similarity between the responses of GPT-4V and human answers as a continuous and categorical variable, respectively.

### Statistical analysis

Statistical analysis was performed using Stata software (version 17.0, StataCorp, College Station, Texas, USA). Stacked bars were graphed using GraphPad Prism (version 9.5.0, GraphPad Software Inc. San Diego, CA, USA).

## Results

### Overall performance

**Table 1** summarizes the performance of GPT-4V in terms of accuracy, usability, and safety based on the proportion of responses rated as good (score of 1) across 600 questions related to ophthalmic multimodal images. Fleiss’ Kappa values indicated good consistency among the three raters, with 0.758 for accuracy, 0.751 for usability, and 0.767 for safety. Only 30.6% (183.3/600) of responses were considered correct by ophthalmologists and 21.5% (123.3/540) were deemed highly usable, while approximately half of the answers (55.6% (300.3/540)) were considered no harm (**Figure 2**).

**Figure 2.**
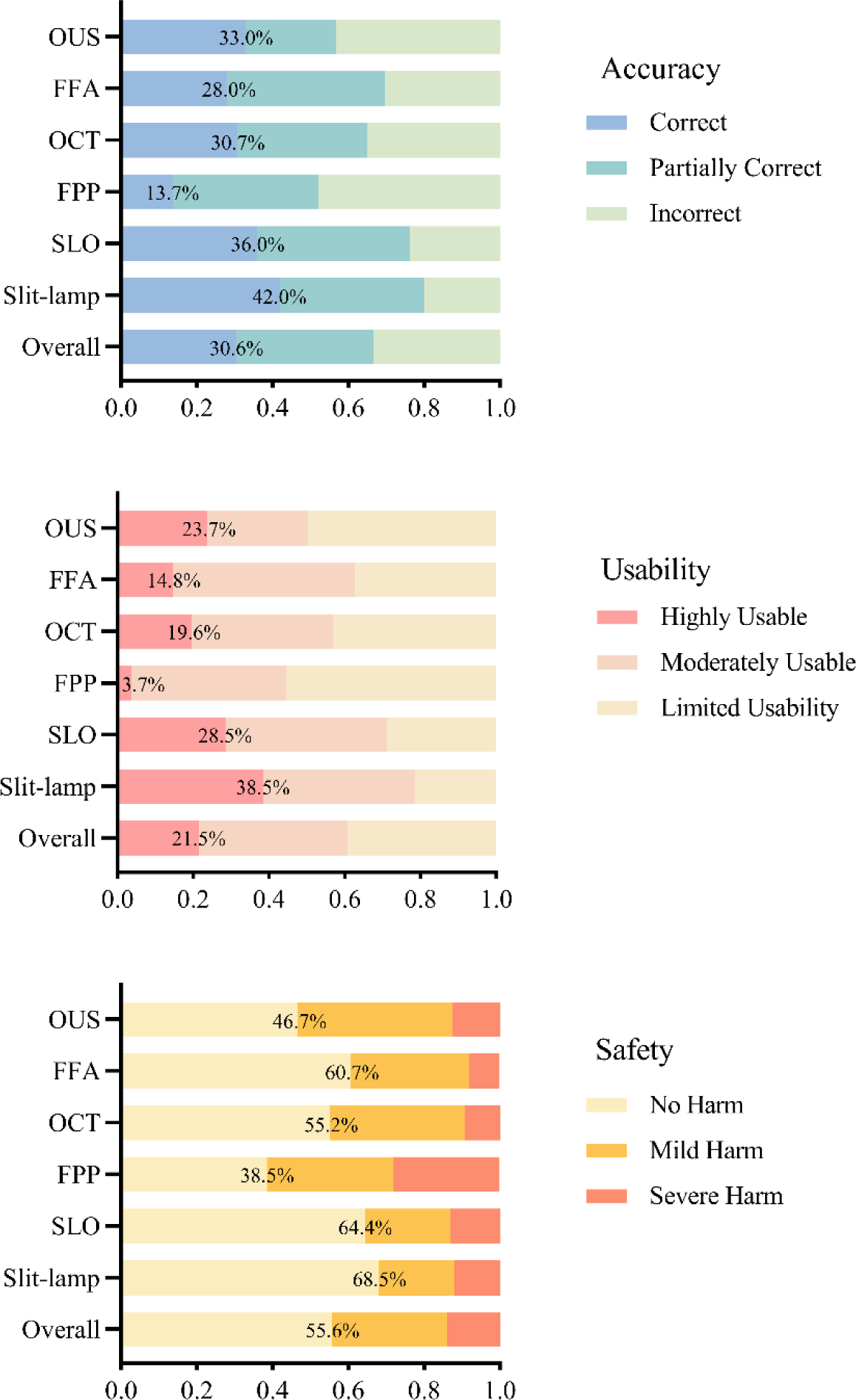
Stacked Bars of Accuracy, Usability and Safety in Ocular Multimodal Images.

**Table 1.** The Intragrader Agreement of three Ophthalmologists in evaluating the Performance of GPT-4V(ision) on Ocular Multimodal Images (600 QA pairs)

|  | Rater 1, N (%)<sup>†</sup> | Rater 2, N (%) | Rater 3, N (%) | Kappa |
| --- | --- | --- | --- | --- |
| **Accuracy** | 179 (29.8) | 183 (30.5) | 187 (31.2) | 0.758 |
| **Usability** | 120 (22.2) | 123 (22.7) | 105 (19.4) | 0.751 |
| **Safety** | 302 (55.9) | 304 (56.3) | 293 (54.3) | 0.767 |

<sup>†</sup> Number and percentage of Correct/Highly Usable/No Harm were presented.

### Multimodal performance

GPT-4V performed best in slit-lamp images, with average proportions of correct, highly usable, and no harm answers being 42.0% (42.0/100), 38.5% (34.7/90), and 68.5% (61.7/90), respectively (**Figure 2**). However, for FPP, only 13.7% (13.7/100), 3.7% (3.3/90), and 38.5% (34.7/90) of answers were rated as correct, highly usable, and no harm, respectively. In contrast, GPT-4V received relatively better evaluations (36.0% correct, 28.5% highly usable, and 64.4% no harm) with SLO images. For detailed evaluations of the six modalities, please refer to **Supplemental Table 4**. The rates of overlap for responses that were correct, highly usable, and no harm were 32.2% (29/90), 23.3% (21/90), 2.2% (2/90), 14.4% (13/90), 8.9% (8/90), and 20.0% (18/90) for slit-lamp, SLO, FPP, OCT, FFA, and OUS, respectively. Overall, only 16.8% (91/540) of responses for the six modalities were correct, highly usable, and no harm.

### Image interpretation performance

We assessed the ability of GPT-4V to interpret ophthalmic images using various VQA tasks, including examination identification, lesion recognition, diagnostic capacity, and decision support (see **Table 2**). Overall, GPT-4V was able to correctly identify the imaging modality of most ophthalmic images (average 95.6% (57.3/60)).

**Table 2.** Evaluation of Identification, Diagnosis, and Reasoning Capabilities of GPT4-V(ision)

|  | Examination<br>Identification<br>(N = 60) | Lesion<br>Identification<br>(N = 60) | Diagnosis<br>Capacity<br>(N = 60) | Decision<br>Support<br>(N = 420) |
| --- | --- | --- | --- | --- |
| **Accuracy** |  |  |  |  |
| Correct, N (%)<sup>†</sup> | 57.3 (95.6) | 15.3 (25.6) | 9.7 (16.1) | 101.0 (24.0) |
| Partially Correct, N (%) | 2.7 (4.4) | 28.3 (47.2) | 14.3 (23.9) | 171.0 (40.7) |
| Incorrect, N (%) | 0 (0) | 16.3 (27.2) | 36.0 (60.0) | 148.0 (35.2) |
| **Usability** |  |  |  |  |
| Highly Usable, N (%) | - | 14.0 (23.3) | 9.0 (15.0) | 93.0 (22.1) |
| Moderately Usable, N (%) | - | 26.0 (43.3) | 14.3 (23.8) | 171.3 (40.8) |
| Limited Usability, N (%) | - | 20.0 (33.3) | 36.7 (61.1) | 155.7 (37.1) |
| **Safety** |  |  |  |  |
| No Harm, N (%) | - | 32.3 (53.9) | 18.7 (31.1) | 249.3 (59.4) |
| Mild Harm, N (%) | - | 18.3 (30.6) | 19.7 (32.8) | 127.0 (30.2) |
| Severe Harm, N (%) | - | 9.3 (15.6) | 21.7 (36.1) | 43.7 (10.4) |

<sup>†</sup> N (%) showed in this table was the mean value of 3 raters.

#### Lesion identification

An average of 25.6% (15.3/60) of answers were considered correct, 47.2% (28.3/60) partially correct, and 27.2% (16.3/60) incorrect. In terms of usability, an average of 23.3% (14.0/60) of answers provided useful recognition of the image’s lesions, 43.3% (26.0/60) were moderately usable, and 33.3% (20.0/60) had limited usability. Regarding potential harm caused by the responses, less than half (46.1% (27.7/60)) were considered unsafe, with 15.6% (9.3/60) deemed to have serious potential harm.

#### Diagnosis capacity

Disease staging and classification were not necessary for this evaluation. If the response achieved top-1 accuracy, meaning that the prediction with the highest probability matched the expected answer exactly,[21] it was considered “correct” during evaluation. If the response achieved top-5 accuracy, which means that the expected answer matched any of the top-5 highest probability predictions, it was assessed as “partially correct”. Otherwise, it was recorded as “incorrect”. The results showed that only 16.1% (9.7/60), 15.0% (9.0/60), and 31.1% (18.7/60) of the responses met the standards of clinical doctors in terms of accuracy, usability, and safety, respectively.

#### Decision support

This involves a range of topics including further examinations, treatment options, visual impact, complications, etiology, prognosis, and prevention measures, requiring the model to have the ability to integrate image information and professional knowledge. We established a relevant series of questions (**Supplemental Table 2**) to test this. The results showed that GPT-4V only met clinical standards (score of 1) in terms of accuracy and usability in 24.0% (101.0/420) and 22.1% (93.0/420) of responses, respectively, while 59.4% (249.3/420) of the responses were considered no harm.

### Evaluation of Repeatability in Diagnosis

As shown in **Supplemental Table 5**, the responses of GPT-4V had the highest repeatability in FFA (100%, 10/10) and the lowest repeatability in OCT (40%, 4/10). The overall repeatability in the diagnosis of six ocular modality images was 63.3% (38/60).

### Sentence Similarity

The overall sentence similarity between the responses of GPT-4V and human answers was 55.5% (**Supplemental Table 6**). Slit-lamp images got the highest score in sentence similarity (61.9%), while FFP images got the lowest (52.3%).

### GPT-4-based Auto Evaluation

This benchmark achieved a Spearman correlation of 0.569 in accuracy and 0.576 in usability using the GPT-4-based auto evaluation. While the Kendall-Tau correlation was 0.456 in accuracy and 0.474 in usability (**Table 3**).

**Table 3.** Auto-evaluation by GPT-based method.

|  | Human<br>Evaluation | GPT-4-based Auto<br>Evaluation* | Spearman<br>correlation | Kendall-Tau<br>correlation |
| --- | --- | --- | --- | --- |
| **Accuracy** |  |  | 0.569 | 0.456 |
| Correct, %<sup>†</sup> | 30.6 | 15.9 | - | - |
| Partially Correct, % | 36.1 | 33.9 | - | - |
| Incorrect, % | 33.4 | 50.2 | - | - |
| **Usability** |  |  | 0.576 | 0.474 |
| Highly Usable, % | 21.5 | 12.4 | - | - |
| Moderately Usable, % | 39.2 | 25.6 | - | - |
| Limited Usability, % | 39.3 | 62.0 | - | - |

<sup>†</sup> The percentage shown in this table represents the average value of 3 raters in human evaluation and the average value of 20 rounds of auto-evaluation based on GPT-4. \* The responses of GPT-4V and human were auto-rated by GPT-4-0613 using the same evaluation criteria in regards to accuracy and usability, and the resulting ratings were compared using the Spearman and Kendall-Tau correlations.

## Discussion

In this study, we evaluated the capacity of GPT-4V to recognize, interpret, and make inferences based on ophthalmic multimodal images. While GPT-4V demonstrated a high level of performance in identifying the modality of ophthalmic images, its ability to recognize lesions, provide diagnoses, and offer decision support was found to be limited. The repeatability in the diagnosis of ophthalmic multimodal images was only 63.3%. The automatic evaluation performance for open-ended questions is still poor in this benchmark. Despite its promising advancements, GPT-4V is not yet ready to support the generation of clinical decisions and patient consultations based on ophthalmic examination images in real-world settings. Importantly, the data used in this study are freely available for other researchers to leverage in their validation or training/fine-tuning experiments.

GPT-4V currently is not ready for clinical use, as only 16.8% of responses were correct, highly usable, and no harm. The preprint study by Wu and colleagues demonstrated GPT-4V’s ability to accurately identify different imaging modalities such as X-ray, CT, MRI, ultrasound, nuclear imaging, and pathology.[10] Our results on ophthalmic multimodal images align with these findings. Out of the 60 images we tested, only three slit-lamp images were identified as close-up photographs of the eye. Notably, two out of these three were diffuse illumination images without the presence of a slit beam, making this classification arguably acceptable. GPT-4V demonstrated varying levels of competency across different imaging modalities. In terms of accuracy, usability, and safety, both slit-lamp and SLO images were rated above average. The high performance on slit-lamp images could be attributable to the similarity these images bear to real-world visual observations, coupled with GPT-4V’s pre-training on disease-specific ophthalmic datasets. GPT-4V’s performance on FPP images was notably below average. Despite both FPP and SLO providing a direct visualization of the fundus, the former only captures a 45-degree field of view of the posterior region. This limited coverage might not fully reveal all pathological features, which could potentially hinder GPT-4V’s ability to accurately recognize and interpret these images. For example, GPT-4V could detect a retinal detachment in SLO but not in FPP. Moreover, in our limited cases, GPT-4V produced several inaccurate descriptions of lesions. For instance, it mischaracterized a typical CRVO image in SLO as pigment changes and misdescribed extensive subretinal exudation in an FPP image of Coats disease as “extensive areas of opacification and discoloration in the retina.” An OCT image of a macular hole without retinal detachment was incorrectly described as “subretinal fluid accumulation and macular detachment”. In a slit-lamp image, the location of an eyelid nevus was erroneously identified as “on the sclera near the limbus”. These inaccuracies underscore that GPT-4V’s recognition of ophthalmic image anatomy and disease characteristics is not reliable. Notably, GPT-4V demonstrated a tendency to provide non-specific, albeit accurate, descriptions of lesions in the evaluation of FFA images. For example, it often generated responses such as, “The images show areas of hyperfluorescence which may suggest leakage, staining, pooling, or abnormal vessel growth, and hypofluorescence which may indicate blockage or non-perfusion.” While this statement is generally correct for any FFA image as it essentially enumerates all possibilities of hyperfluorescence and hypofluorescence, it does not provide a specific analysis for the presented images. We therefore conclude that GPT-4V does not currently possess the ability to analyze the specific localization of lesions in FFA images. It instead tends to produce generic responses that could be perceived as “boilerplate” answers. The significantly higher accuracy (70%) achieved by Mihalache A et al. using GPT-4V on OCT images might be attributed to their use of multiple-choice questions, while our study used open-ended questions. [22]

The repeatability of GPT-4V in diagnosing ocular images was low, with only 63.3% (38/60) of the responses being consistent. GPT-4V showed the highest repeatability on FFA, but most of its responses were vague and generic. For example, it often gave answers like “the patterns could suggest a retinal vascular disorder such as diabetic retinopathy, retinal vein occlusion, or choroidal neovascularization from conditions like wet age-related macular degeneration”. These answers did not specify the exact diagnosis or the features that supported it. Therefore, GPT-4V performed poorly in accuracy.

The automatic evaluation performance for open-ended questions is still poor in this benchmark. Recently, automatic evaluation for Open-QA has got significant progress, especially with the use of LLM.[19 23] In this study, we got Spearman and Kendall-Tau correlation results similar to previous study,[20] which also indicates that automatic evaluation for Open-QA remains a major challenge today and there is still a significant research space for improvement. The reason we did not conduct an automated evaluation in terms of safety is that automated evaluation has not yet reached a level of trustworthiness comparable to human evaluation.

As a pioneering evaluation of GPT-4V’s capabilities in processing ophthalmic images, our study adds valuable insights to the existing body of knowledge. In medical scenarios, GPT-4V appears to have a strong safeguard system in place to prevent it from making direct diagnoses.[10] In our study, when faced with relatively rare ophthalmic conditions, GPT-4V typically refrained from making explicit diagnoses, instead suggesting a few potential diseases. However, upon evaluation by clinical doctors, the diseases it suggested did not resemble the ground truth given by ophthalmologists. Consequently, its subsequent clinical decision-making suggestions, based on inaccurate or imprecise diagnoses, were unreliable. A model exhibiting higher performance may assist clinical doctors in decision support, interpreting medical conditions, and providing high-quality healthcare consultation in remote areas. One promising way of improving multimodal models is to build foundation models with self-supervised pretraining with a large number of retinal images.[24] Another way to improve the model is to align the ophthalmic image features with medical texts.[11 25] Additionally, researchers may improve their models by applying transfer learning, fine-tuning, and reinforcement learning techniques, or by incorporating knowledge enrichment methods like Retrieval-Augmented Generation (RAG).[26 27] However, before applying any model to clinical practice, detailed clinical trials should be conducted to assess its safety and efficacy. Even if ultimately used in clinical settings, it also should be used under the supervision of human clinical doctors. This study provides a benchmark for further investigation in building large language models for processing ophthalmic multimodal images.

There are several limitations in our study. Firstly, we only assessed ten selected images for each specific modality, which may introduce a sample bias. Future studies could benefit from expanding and diversifying the dataset to better represent the variability seen in clinical practice. Currently, evaluation methods for the VQA capabilities of multi-modal large language models like GPT-4V are limited, especially for knowledge-intensive scenarios.[9 28] The establishment of benchmarks for these scenarios has yet to reach a consensus.[29] Secondly, we did not attempt to further prompt GPT-4V in instances where its responses were incorrect, thereby not testing its self-correction abilities. Exploring this aspect could be an interesting avenue for future research. Thirdly, the dataset only covers 60 ophthalmic conditions and may not fully represent the diversity of real-world clinical situations. Nevertheless, it includes a range of common ophthalmic conditions, which can provide a benchmark for assessing the capabilities of LLM.

Overall, GPT-4V is not yet ready to generate clinical decisions or provide patient consultations based on images from ophthalmic examinations in real-world scenarios. Ongoing refinement and testing remain crucial to enhance the performance of large language models in ophthalmology.

## Contributors

PX, XC, ZZ and DS designed the study; DS provided data. PX, XC and ZZ analysed and interpreted the data; PX, XC and ZZ drafted the manuscript; DS revised the manuscript. DS is responsible for the overall content as the guarantor.

## Patient consent for publication

Not applicable.

## Ethics approval

This study was approved by the institutional review board of The Hong Kong Polytechnic University.

## Provenance and peer review

Not commissioned; externally peer-reviewed.

## Data availability statement

The OphthalVQA dataset will be freely accessible on figshare (the link will be provided upon article acceptance).

---

**Supplemental Figure 1. Screenshot of the process of building a digital ophthalmologist based on GPT-4V(ision).**

