## Supplemental Table 1 for "Unveiling the Clinical Incapabilities: A Benchmarking Study of GPT-4V(ision) for Ophthalmic Multimodal Image Analysis"

**Supplemental Table 1. Diseases Category in the Evaluation Dataset.**

| Modality | Diagnosis |
|---|---|
| Slit-lamp | Cataract, pterygium, subconjunctival hemorrhage, conjunctivitis, keratitis, corneal conjunctivalization, intraocular lens dislocation, corneal dermoid, anterior uveitis, and eyelid mass |
| SLO | Central retinal vein occlusion, proliferative diabetic retinopathy, rhegmatogenous retinal detachment, macular hole, pathologic myopia, central serous chorioretinopathy, retinitis pigmentosa, branch retinal artery occlusion, choroidal coloboma, and Coat's disease |
| FPP | High myopia, branch retinal vein occlusion, recent retinal detachment, nonproliferative diabetic retinopathy, chronic tractional retinal detachment, Coat's disease, choroidal coloboma, intraocular lens dislocation, myelinated retinal nerve fiber, and choroidal melanoma |
| OCT | Macular hole, macular epiretinal membrane, cystoid macular edema, retinal detachment, age-related macular degeneration, polypoidal choroidal vasculopathy, central serous chorioretinopathy, choroidal neovascularization, myopic foveoschisis, and retinal pigment epithelial detachments |
| FFA | Proliferative diabetic retinopathy, central retinal vein occlusion, central retinal artery occlusion, age-related macular degeneration, pathologic myopia, Coat's disease, central serous chorioretinopathy, retinitis pigmentosa, familial exudative vitreoretinopathy, and intermediate uveitis |
| OUS | Vitreous opacities, silicone oil-filled eye with silicone oil emulsification, retinal detachment, choroidal detachment, high myopia with posterior scleral staphyloma, Tenon's capsule edema, foreign body in the vitreous cavity, solid orbital mass, solid intraocular mass with retinal detachment, and retinoblastoma |

SLO = Scanning Laser Ophthalmoscopy; FPP = Fundus Photography of the Posterior Pole; OCT = Optical Coherence Tomography; FFA = Fundus Fluorescein Angiography; OUS = Ocular Ultrasound.
