## Supplemental Table 2 for "Unveiling the Clinical Incapabilities: A Benchmarking Study of GPT-4V(ision) for Ophthalmic Multimodal Image Analysis"

**Supplemental Table 2. Question Lists.**

| Capability | Questions |
|---|---|
| Examination Identification | What kind of examination is this image? |
| Lesion Identification | Is there any abnormality? |
| Diagnosis capacity | What is the diagnosis? |
| | What examinations need to be done next? |
| | What might be the treatment options for this condition? |
| | Can this condition cause blindness? |
| Decision Support | What are some common complications that can occur? |
| | What might be the cause of this condition? |
| | Will this condition progress? |
| | How can I prevent this condition? |
