## Supplemental Table 3 for "Unveiling the Clinical Incapabilities: A Benchmarking Study of GPT-4V(ision) for Ophthalmic Multimodal Image Analysis"

**Supplemental Table 3. Example Question-Answer on Ocular Images Using GPT-4V(ision).**

| Image | Question | Answer of GPT-4V |
|---|---|---|
| 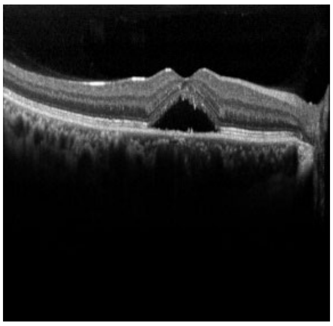 | What kind of examination is this image? | This image is from an Optical Coherence Tomography (OCT) examination, which is commonly used to capture high-resolution cross-sectional images of the retina. |
| | Is there any abnormality? | Yes, there is an abnormality. The image shows a small, dark, dome-shaped area in the central retina which is consistent with fluid accumulation beneath the retinal layers. |
| | What is the diagnosis? | The diagnosis suggested by this OCT image could be central serous chorioretinopathy (CSR), which is characterized by the accumulation of subretinal fluid in the macula. |
| | What examinations need to be done next? | Additional examinations could include a fluorescein angiography (FA) to detect any leaking areas that may be contributing to the fluid accumulation and to observe the overall retinal circulation. An Amsler grid test may also be beneficial to determine the extent of central visual field distortion. |
| | What might be the treatment options for this condition? | Treatment for CSR often begins with observation as many cases resolve on their own. If treatment is necessary, options may include laser therapy or photodynamic therapy to reduce leakage and promote fluid resorption. |
| | Can this condition cause blindness? | CSR typically affects central vision and can lead to significant vision impairment if chronic or recurrent. However, it does not usually result in complete blindness as peripheral vision is generally not affected. |
| | What are some common complications that can occur? | Common complications from CSR can include persistent central vision loss, recurrent episodes, and in some cases, permanent retinal changes that may affect vision long-term. |
| | What might be the cause of this condition? | The cause of CSR is not fully understood, but it is thought to be associated with stress, corticosteroid use, and sometimes Type A personality traits. |
| | Will this condition progress? | CSR can progress, with some cases resolving spontaneously within months, while others may develop chronic or recurrent fluid accumulation. |
| | How can I prevent this condition? | Preventing CSR involves managing stress, avoiding corticosteroid use if possible, and addressing any associated systemic health issues. Regular eye exams are essential for individuals with risk factors for early detection and management. |
