## Supplemental Table 4 for "Unveiling the Clinical Incapabilities: A Benchmarking Study of GPT-4V(ision) for Ophthalmic Multimodal Image Analysis"

**Supplemental Table 4. Evaluation of the Performance of GPT-4V(ision) in Ocular Multimodal Images**

| | Overall | Slit-lamp | SLO | FPP | OCT | FFA | OUS |
|---|---|---|---|---|---|---|---|
| **Accuracy** | | | | | | | |
| Correct, N(%)† | 183.3(30.6) | 42.0(42.0) | 36.0(36.0) | 13.7(13.7) | 30.7(30.7) | 28.0(28.0) | 33.0(33.0) |
| Partially Correct, N(%) | 216.3(36.1) | 38.0(38.0) | 40.3(40.3) | 38.3(38.3) | 34.3(34.3) | 41.7(41.7) | 23.7(23.7) |
| Incorrect, N(%) | 200.3(33.4) | 20.0(20.0) | 23.7(23.7) | 48.0(48.0) | 35.0(35.0) | 30.3(30.3) | 43.3(43.3) |
| **Usability** | | | | | | | |
| Highly Usable, N(%) | 116.0(21.5) | 34.7(38.5) | 25.7(28.5) | 3.3(3.7) | 17.7(19.6) | 13.3(14.8) | 21.3(23.7) |
| Moderately Usable, N(%) | 211.7(39.2) | 36.0(40.0) | 38.3(42.6) | 36.7(40.7) | 33.7(37.4) | 43.0(47.8) | 24.0(26.7) |
| Limited Usability, N(%) | 212.3(39.3) | 19.3(21.5) | 26.0(28.9) | 50.0(55.6) | 38.7(43.0) | 33.7(37.4) | 44.7(49.6) |
| **Safety** | | | | | | | |
| No Harm, N(%) | 300.3(55.6) | 61.7(68.5) | 58.0(64.4) | 34.7(38.5) | 49.7(55.2) | 54.7(60.7) | 42.0(46.7) |
| Mild Harm, N(%) | 165.0(30.6) | 17.7(19.6) | 20.3(22.6) | 30.0(33.3) | 32.0(35.6) | 28.0(31.1) | 36.7(40.7) |
| Severe Harm, N(%) | 74.7(13.8) | 10.7(11.9) | 11.7(13.0) | 25.3(28.1) | 8.3(9.3) | 7.3(8.1) | 11.3(12.6) |

† N (%) showed in this table was the mean value of 3 raters. SLO = Scanning Laser Ophthalmoscopy; FPP = Fundus Photography of the Posterior Pole; OCT = Optical Coherence Tomography; FFA = Fundus Fluorescein Angiography; OUS = Ocular Ultrasound.
