## Supplemental Table 5 for "Unveiling the Clinical Incapabilities: A Benchmarking Study of GPT-4V(ision) for Ophthalmic Multimodal Image Analysis"

**Supplemental Table 5. Evaluation of Repeatability in Diagnosis**

| Modality | Repeatability, %(consistent response/total) |
|---|---|
| Slit-lamp | 70.0 (7/10) |
| SLO | 50.0 (5/10) |
| FPP | 70.0 (7/10) |
| OCT | 40.0 (4/10) |
| FFA | 100.0 (10/10) |
| OUS | 50.0 (5/10) |
| Overall | 63.3 (38/60) |

SLO = Scanning Laser Ophthalmoscopy; FPP = Fundus Photography of the Posterior Pole; OCT = Optical Coherence Tomography; FFA = Fundus Fluorescein Angiography; OUS = Ocular Ultrasound.
