## Supplemental Table 6 for "Unveiling the Clinical Incapabilities: A Benchmarking Study of GPT-4V(ision) for Ophthalmic Multimodal Image Analysis"

**Supplemental Table 6. Auto-evaluation by Sentence Similarity Between the Response of GPT-4V and Human Answer.**

| Modality | Sentence Similarity, % |
|---|---|
| Slit-lamp | 61.9 |
| SLO | 52.7 |
| FPP | 52.3 |
| OCT | 56.7 |
| FFA | 56.1 |
| OUS | 53.3 |
| Overall | 55.5 |

SLO = Scanning Laser Ophthalmoscopy; FPP = Fundus Photography of the Posterior Pole; OCT = Optical Coherence Tomography; FFA = Fundus Fluorescein Angiography; OUS = Ocular Ultrasound.
