## Supplemental Figure 1 for "Unveiling the Clinical Incapabilities: A Benchmarking Study of GPT-4V(ision) for Ophthalmic Multimodal Image Analysis"

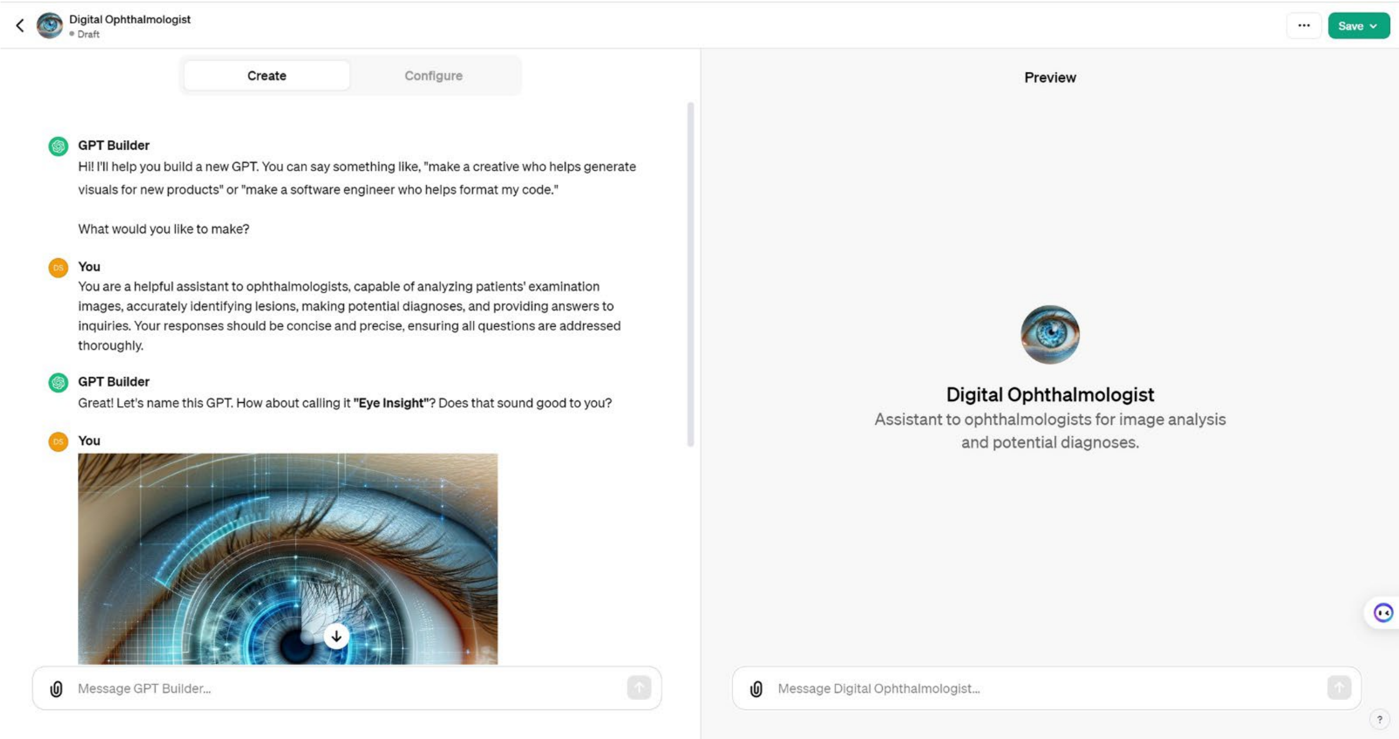


**Supplemental Figure 1. Screenshot of the process of building a digital ophthalmologist based on GPT-4V(ision).**
